# Evaluating whole fruit feijoa powder for type 2 diabetes risk prevention: the FERDINAND RCT

**DOI:** 10.1101/2025.06.18.25329000

**Authors:** I.A. Mohamed, I.R. Sequeira-Bisson, J.J. Lim, L.D. Plank, R. Murphy, S.D. Poppitt, J.L. Miles-Chan

**Author notes:** These authors share senior authorship (S.D. Poppitt, J.L. Miles-Chan). **Corresponding Author:**Prof Jennifer Miles-Chan, Human Nutrition Unit, 18 Carrick Place, Mount Eden, Auckland 1024, New Zealand.

## Abstract

**Background:** Low energy diets (LEDs) are effective for body weight (BW) loss and improvement of type 2 diabetes (T2D) risk biomarkers. Emerging evidence suggests that whole fruit feijoa powder, rich in polyphenols and abscisic acid, may further support T2D prevention. The FERDINAND study investigated whether daily consumption of a whole fruit feijoa powder enhanced LED-driven improvements in fasting plasma glucose (FPG), BW, and other metabolic markers.

**Methods:** At in-clinic screening, 97 participants were enrolled into the trial and randomised to receive 1.15 g/day of whole fruit feijoa powder (Fx, n=48) or Placebo treatment (n=49) for 6 months. All participants underwent 2 months of LED-induced weight loss, followed by 4 months of dietary advice for weight loss maintenance. BW, FPG, and secondary outcomes including blood pressure (BP), were assessed at baseline (M0), end of month 2 (M2), 4 (M4), and 6 (M6). Data were analysed using linear mixed-effects models as intention-to-treat (ITT) with imputation for missing not-at-random data points.

**Results:** BW significantly decreased in both treatment groups during the 2 month LED, with a gradual upward trajectory over the subsequent 4 months (time_0-6m_: P<0.001). There was no difference in BW between Fx and Placebo over the 6 month intervention (treatment×time_0-6m_: P=0.74). FPG followed the weight loss trajectory in both treatment groups (time_0-6m_: P<0.001), but with no significant between-group interaction over 6 months (treatment×time_0-6m_: P=0.09, n=97). Systolic BP (SBP), but not diastolic BP, declined significantly over the 6 months between treatments (treatment×time_0-6m_: P=0.01) in all participants, with SBP in Fx lower than Placebo at M4 and M6 (P<0.05, both).

**Conclusions:** In adults with overweight and prediabetes, Fx supplementation may enhance blood pressure improvement achieved through BW loss. The polyphenolic matrix of whole fruit feijoa, which contains a high proportion of catechins, may have contributed to vascular benefits.

## Introduction

Type 2 diabetes (T2D), which comprises 90-95 % of all diabetes cases (1), has become an ever-burgeoning strain on global health. An estimated 589 million adults worldwide currently have diabetes, with a 45% increase in cases projected by 2050 (2). In the last two decades, there has been a three-fold increase in T2D-related global health expenditure, which now exceeds USD 1 trillion (2). The number of individuals with prediabetes is even greater. Prediabetes is a state of dysglycaemia, whereby blood glucose levels are elevated above normal cut-offs, but below threshold for diabetes diagnosis. Over 1 billion adults worldwide have prediabetes, with cases expected to approach 1.5 billion by 2050 (2). Therefore, implementing targeted T2D prevention, especially during prediabetes, is critical in alleviating global health burden.

Extensive evidence points to excess adiposity being the biggest risk factor for T2D (3–10). This coupled with the increasing prevalence of obesity, makes body weight (BW) loss key in T2D prevention. Bariatric surgery and pharmaceutical agents are widely used for BW loss. However, these tools come with several drawbacks, such as limited availability and prohibitive costs. A less invasive method for achieving BW loss is through commercial low or very low energy diets (LED or VLED), implemented using nutritionally complete meal replacement products (11–13). LEDs encompass a daily energy intake between 3347-5021 kJ, while energy intake ≤3347 kJ characterises VLEDs (12,14). A large body of evidence shows that LEDs and VLEDs, combined with behavioural support, are effective for BW loss and improving T2D biomarkers (15–23).

Dietary composition plays an important role in T2D, with risk reduction associated with lowered refined grain intake and increased intake of whole grains, fruits, vegetables, legumes, and nuts (24–26). Consequently, identifying foods which may enhance proven strategies, such as LEDs, is of great importance. Feijoa (*Acca sellowiana*), also known as pineapple guava, is a fruit originating in South America and widely grown in New Zealand (27,28), with biochemical properties of promise in lowering T2D risk. Specifically, it is abundant in polyphenols and abscisic acid (ABA, a plant compound named for its role in abscission), particularly in the skin (29,30). Evidence of polyphenols eliciting T2D risk reduction has been well established (31–42), with evidence for ABA emerging recently (43–46). Mechanistically, ABA exhibits binding to pertussis toxin-sensitive G protein-coupled receptor (GPCR) (47), peroxisome proliferator-activated receptor gamma (PPAR-γ) (48), and lanthionine synthetase C-like protein 2 (LANCL2) (49,50). Activation of these targets promote intracellular signalling cascades involving cyclic adenosine monophosphate, cyclic adenosine diphosphate ribose, and protein kinase B phosphorylation, which enhance insulin secretion, glucose uptake, and translocation of glucose transporter type 4 (GLUT4) to the plasma membrane (51–56).

To-date, only 2 studies (1 pre-clinical, 1 clinical) have investigated the effect of feijoa on BW and T2D biomarkers. In leptin deficient ob/ob mice, intake of feijoa extract (300 mg/kg BW/day) significantly decreased BW gain, liver organ weight, and circulating total cholesterol (57). In a small sample randomised controlled trial (RCT) of 34 adults with T2D, conducted in Iran, 150 mg/day of feijoa extract for 12 weeks significantly lowered fasting plasma glucose (FPG), glycated haemoglobin (HbA_1c_), and triglycerides (TAG), compared to placebo (58). We hypothesised that whole fruit feijoa powder when combined with meal replacement LED, would produce greater benefits than LED alone on acute and/or longer-term weight loss or weight loss maintenance, along with associated glycaemic and other metabolic outcomes. Therefore, the 6-month FERDINAND (evaluating <u>FE</u>ijoa fo<u>R Di</u>abetes prevention in a multi-ethnic <u>N</u>ew Ze<u>A</u>la<u>ND</u> cohort) study investigated daily consumption of 1150 mg/day whole fruit feijoa powder during 2-months of LED-driven weight loss followed by 4-months of weight maintenance, in a multiethnic New Zealand cohort with overweight and prediabetes.

## Methods

### Study Design

The FERDINAND study was a two-arm, parallel design, double-blind RCT, of 6 months duration (Figure 1). Participants underwent 2-months of LED-driven weight loss (i.e. LED phase) followed by 4-months of weight loss maintenance (i.e. maintenance phase). Participants consumed either whole fruit feijoa powder or Placebo powder every day for the entire study period. The whole fruit feijoa powder consisted of 1,150 mg of microcrystalline cellulose and 1,150 mg of freeze-dried powder with skin included (Feiolix®, Anagenix Ltd, Auckland, New Zealand; Fx). The Placebo powder sachet contained 2,300 mg of microcrystalline cellulose. The study received ethical approval from the New Zealand Health and Disabilities Committee (Reference: 2022 EXP 12032) and was also prospectively registered with the Australian New Zealand Clinical Trials Registry (ACTRN: 12622000210774). The study was conducted at the Human Nutrition Unit (HNU), University of Auckland, New Zealand, from September 2022 to March 2024.

**Figure 1:**
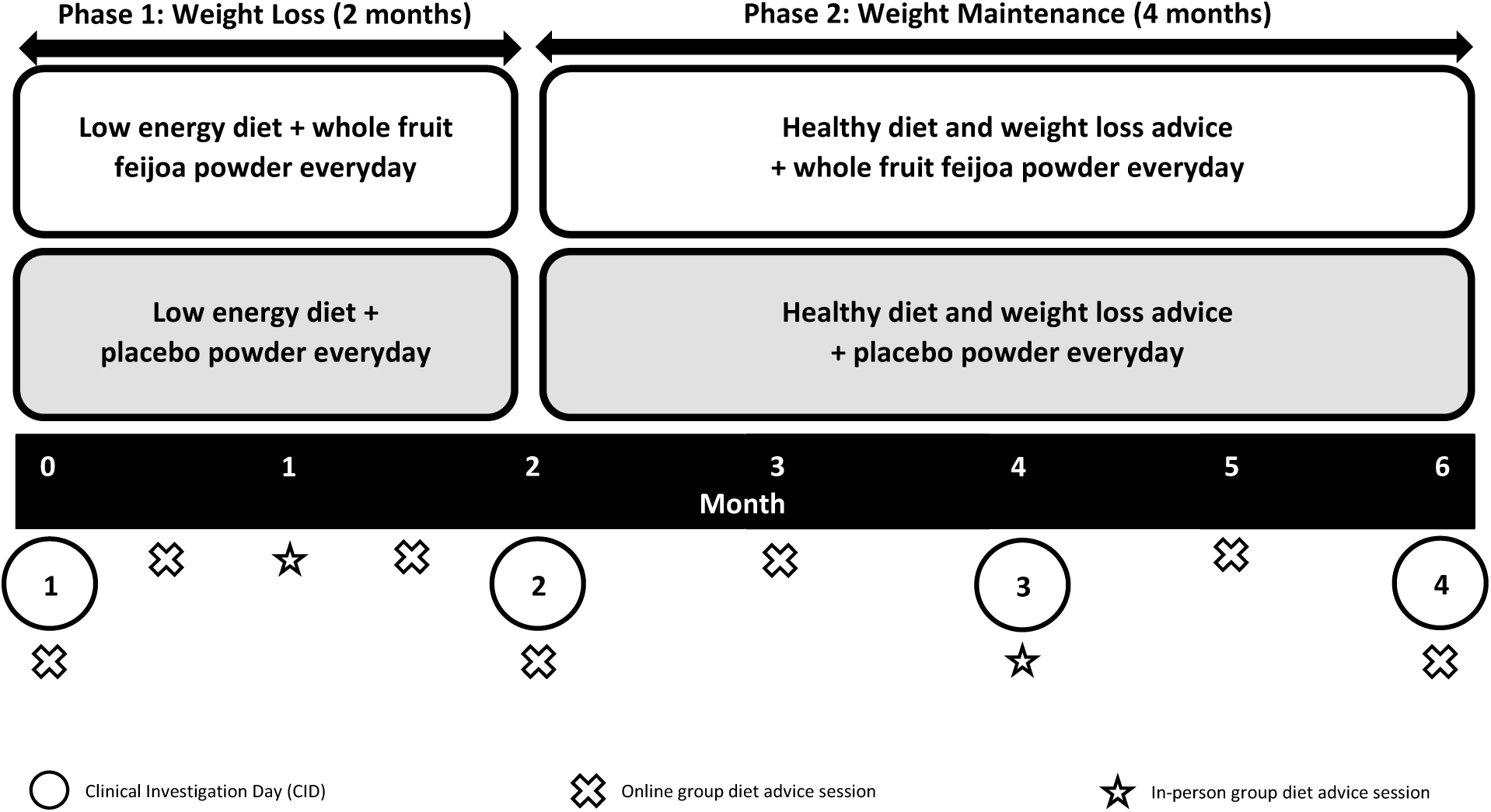
FERDINAND Study Design.

### Participants

Prospective participants living in the wider Auckland area were pre-screened via an online questionnaire and telephone interview, then invited to the HNU for an on-site screening visit. Prior to any data collection, individuals provided written informed consent. Prediabetes status was determined by FPG, using American Diabetes Association guidelines (ADA, 5.6-6.9 mmol/L) (1). Hypertension was defined as SBP ≥140 mmHg and/or DBP ≥90 mmHg (uncontrolled hypertension) and/or use of antihypertensive medications (controlled hypertension) (59,60). The other inclusion criteria were body mass index (BMI) ≥26 kg/m^2^, BW ≤150 kg, Finnish Diabetes Risk Score ≥12, otherwise healthy per self-report and able to attend clinical investigation day (CID) visits. Potential participants were excluded if they had current or history of significant disease, including type 1 or type 2 diabetes mellitus, cardiovascular disease, pancreatic disease, digestive diseases (e.g. inflammatory bowel syndrome/disease, ulcerative colitis, Crohn’s disease) and cancer, recent BW loss/gain >10% within previous three months or were taking part in an active diet program, previous bariatric surgery, current medication controlling glycaemia and weight loss, steroid medications (except topical steroids) or atypical antipsychotics within previous three months, were intending to alter physical activity during following twelve months, smoked, vaped or used recreational drugs within previous six months, or were pregnant or breastfeeding within previous six months. Additional exclusions included unwillingness to consume LED meal replacement products or adhere to the food avoidance requirements during the weight maintenance phase or hypersensitivities/allergies to these foods, not prepared to comply with the study protocol, or participation in other clinical intervention studies in the previous six months.

### Randomisation and Blinding

Participants were randomly allocated in a 1:1 ratio to consume either Fx or Placebo powder following a computer-generated randomisation sequence, with the powders distinguished using anonymised codes and blinding conducted by an independent researcher who maintained confidentiality of the codes until the completion of the study. Participants and the study team remained blinded throughout the study period and were unblinded upon completion of data collection. Participants were randomised to treatment and commenced the study in multiple cohorts each consisting of 10-25 eligible participants.

### Procedures

During the LED phase, participants consumed commercial, complete meal replacement products (Cambridge Weight Plan, Corby, UK), whereby 4 energy-controlled sachets, each containing approximately 840 kJ, were consumed every day. Each meal replacement sachet was reconstituted using 250 mL of water only. Furthermore, participants consumed the intervention powder every day with their first LED product of the day. Participants were prohibited from consuming any other foods or beverages (including coffee, tea, and non-caloric beverages apart from water) during the LED phase, to ensure that any observed effects were from Fx, rather than other foods which may contain the same, or additional, bioactive compounds. Participants were also instructed to drink at least 2.25 L water daily. During the LED phase, Benefiber (GlaxoSmithKline, London, UK) was provided upon request, to participants who self-reported constipation and/or bowel discomfort.

Upon completion of the LED phase, weight loss maintenance was monitored for 4 months. Restrictions on food and beverage intake were mostly removed during this maintenance phase, with feijoa and feijoa-containing products being the only prohibited items. Participants continued consuming their assigned intervention powder every day with their breakfast meal. Group diet advice sessions were conducted by registered dietitians at baseline and throughout the study (Figure 1). These sessions focused on behavioural strategies to enhance adherence to study protocol and long-term considerations for weight maintenance (e.g. meal planning, dealing with food cravings and how to navigate social gatherings). The advice adhered to the New Zealand Ministry of Health healthy eating guidelines for improving metabolic health (61). For both intervention powders and meal replacement products, participants were instructed to keep all opened and unopened sachets and return them at their following CID visit.

It was assumed that opened sachets equated to consumed sachets. Participants were instructed to contact the study team if sachets were opened but not consumed. Furthermore, participants were contacted regularly throughout the study by a trained researcher, to discuss meal replacement and intervention powder compliance. The following calculations were used to assess compliance:

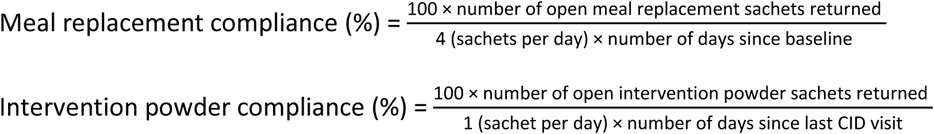

### Outcomes

BW and FPG, were joint co-primary outcomes, while secondary outcomes comprised body composition, blood pressure, and blood biochemistry. All outcomes were assessed at M0/CID1 (baseline), M2/CID2 (post-LED), M4/CID3, and M6/CID4 (study end). Prior to BW, height, and body composition measurements, participants were asked to remove shoes, heavy clothing, headwear (if applicable), jewellery, and all items from their pockets. BW and height were measured in duplicate using a calibrated digital scale (Mettler Toledo Spider 2, Greifensee, Switzerland) and a wall-mounted stadiometer (Seca Model 222, Hamburg, Germany), respectively, and averaged for data analyses. BMI was then calculated using the average of the BW and height measurements. Waist circumference (WC) was measured using a non-stretchable measuring tape (Abbott Laboratories, Illinois, USA), placed horizontally around the abdomen at the level of the umbilicus. Participants inhaled and exhaled once, with the measurement recorded at the end of expiration in duplicate and averaged for data analysis. Body composition was measured, at the University of Auckland Body Composition Laboratory, by dual energy x-ray absorptiometry (DXA) using an iDXA scanner (GE Healthcare, Wisconsin, USA) and analysed using enCORE software (version 18, GE Healthcare, Wisconsin, USA). The following measures were calculated by proprietary algorithms in the enCORE software: total body fat mass (TBFm), total body fat-free mass (TBFFm), abdominal adipose tissue mass (AATm), visceral adipose tissue mass (VATm), and visceral adipose tissue percentage (VATp). Additional measures were also obtained using the following calculations:

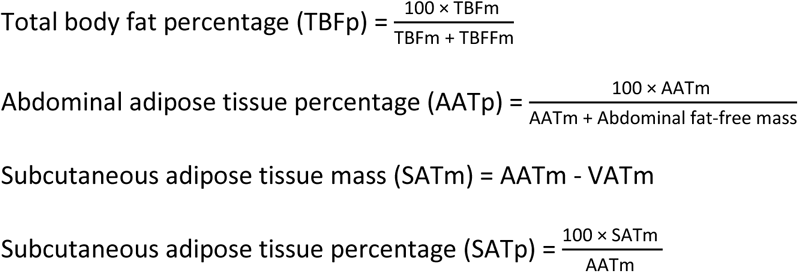

Systolic blood pressure (SBP) and diastolic blood pressure (DBP) were measured in duplicate using an automated sphygmomanometer (Welch Allyn ProBP 2400, Welch Allyn, New York, USA) and averaged for data analyses. Fasting venous blood samples were collected into BD Vacutainer tubes (Becton, Dickinson and Company, New Jersey, USA). Plasma samples were collected for analysis of FPG and glucoregulatory peptides in fluoride oxalate and P800 tubes, respectively. Whole blood was collected in a tripotassium ethylenediaminetetraacetic acid (K_3_-EDTA) tube for HbA_1c_ analysis. Serum samples were collected for lipid biomarkers and liver function enzymes using a serum separator (SST™ ll Advance) tube. The following clinical blood markers were analysed on the cobas® c311 (Roche Diagnostics Ltd, Mannheim, Germany): FPG, HbA_1c_, total cholesterol, TAG, high density lipoprotein cholesterol (HDL-C), low density lipoprotein cholesterol (LDL-C), alanine aminotransferase (ALT), aspartate aminotransferase (AST), alkaline phosphatase (ALP), and gamma-glutamyl transferase (GGT). Insulin and C-peptide were analysed on the cobas® e601 (Roche Diagnostics Ltd, Mannheim, Germany). The online Homeostatic Model Assessment (HOMA) Calculator^©^ (The University of Oxford, version 2.2.3) was used to calculate insulin resistance (HOMA2-IR), β-cell function (HOMA2-B), and insulin sensitivity (HOMA2-S), determined based on FPG and fasting insulin.

### Statistical analysis

Power calculations were conducted based on FPG, using data obtained from the New Zealand arm of the PREVIEW diabetes prevention study (62) conducted in our laboratory in a cohort of adults with overweight and prediabetes, and of mixed ethnicity. The sample size calculations assumed that any FPG improvements achieved during LED phase would be maintained between the treatments during maintenance phase. In a two-arm, parallel-design longitudinal study with baseline FPG of 5.8 mmol/L [standard deviation (SD): 0.6 mmol/L], effect size of 0.5, type I error α=0.05, 80% power, and two-tailed p-value, a clinically significant difference in FPG between treatments of 0.3 mmol/L was expected to be detected as statistically significant in a cohort of 160 participants (80 per treatment), including 20% drop out. Conduct of the trial during the COVID pandemic slowed recruitment however, and re-assessment of sample size showed that a group of n=90 participants (45 per treatment) would still be expected to detect a between-group difference in FPG of 0.3 mmol/L, with SD of baseline FPG 0.5 mmol/L, effect size of 0.6, type I error α=0.05, and power of 80% as significant. Baseline characteristics were summarised using mean and SD for continuous outcomes. Comparison of continuous outcomes, by treatment, were made using Welch’s two-sample t-test. Pearson’s chi-squared test with Yates’ continuity correction was used for comparing categorical outcomes. Attrition rates between the treatments were compared using the likelihood ratio test. In participants who successfully completed the LED phase, compliance to the meal replacements was compared between treatments using Welch’s two-sample t-test. In participants who successfully completed the 6 month intervention, compliance to Fx and Placebo powders was also compared using Welch’s two-sample t-test.

Analyses of all outcomes were conducted as repeated measures linear mixed models (LMM). Participants were included as the random effect. Treatment group (Fx, Placebo), time (M0/CID1, M2/CID2, M4/CID3, M6/CID4), treatment×time interaction, and gender were included as fixed effects, while baseline measurement (M0/CID1) was included as a covariate. When main effect of time was significant, *post hoc* multiple comparisons between the following timepoints were conducted using Tukey’s analyses; M0-M2, M0-M4, M0-M6, M2-M4, M2-M6. To improve normality, the following variables were natural log-transformed prior to conducting LMMs: insulin, HOMA2-IR, TAG, ALT, AST, and GGT. For interpretability, estimated marginal means and SEMs from models involving log-transformed variables were back-transformed by raising them to the power of *e*. All LMM analyses were conducted using intention-to-treat (ITT) approach, for all participants randomised into the trial at M0/CID1, with imputation for missing-not-at-random (MNAR) data points using the last value carried forward (LVCF) method. Given that 80 % of withdrawals occurred during the LED phase (M0-M2), during which metabolic improvements are typically expected even with partial adherence, the ITT strategy provided a conservative estimate of treatment effects. This method is well established in trials of similar design, including the DROPLET (18), DiRECT (22), and DIADEM-I (23) studies.

Noting that Restricted Maximum Likelihood (REML) for LMMs may not adequately handle MNAR data, we also investigated outcomes from the raw data set (no imputation). One extreme FPG outlier (n=1, Placebo; single data point M6/CID4: 9.6 mmol/L, >3 IQR above the upper quartile) was identified, therefore a sensitivity analysis was undertaken where FPG was analysed both with and without this data point. All analyses were conducted using R version 4.3.3 (63). The level of significance was set at P≤0.05.

## Results

### Baseline characteristics, attrition, and compliance

The baseline characteristics of the ITT cohort (n=97) are shown in Table 1. 81.4% of the cohort were female and 18.6% were male. Participants were ethnically diverse, with representation of European Caucasian (45.4%), Māori (17.5%), Pacific Peoples (18.6%), Asian Chinese (12.4%), Asian Indian (9.3%), and Other (16.5%) ethnicities. BW (mean ± SD) was 103.1 ± 21.0 kg and FPG was 6.2 ± 0.6 mmol/L. The percentage of participants who completed the 6 month study in the Fx and Placebo groups was 66.7% and 61.2% respectively. Attrition rates were not significantly different between the groups. Three quarters of withdrawals occurred during the LED weight loss phase (Figure 2). There was 1 adverse event (AE) reported in the Fx group during the LED, whereby the participant experienced intolerance to the meal replacement products. There was also 1 AE reported in the Placebo group during the LED, whereby the participant experienced intolerance to the placebo powder. These participants were withdrawn from the study, upon review by the independent medical monitor. Of those who completed the LED phase, >80% compliance to the meal replacements was achieved in both treatments. Furthermore, the mean ± SD BW loss (M0-M2) in these LED completers was 10.2 ± 4.4 kg, or 10.0 ± 3.5 % (raw data, no imputation). Compliance to Fx and Placebo powders throughout the 6 month study was also >80%, with higher compliance in Placebo (92 ± 11 %) vs Fx (82 ± 22 %; P=0.03).

**Table 1:** Baseline characteristics of ITT cohort at M0/CID1, in all participants and by treatment.

|  | All participants (n=97) | Fx (n=48) | Placebo (n=49) |
| --- | --- | --- | --- |
| **Demographics** |  |  |  |
| Age (yrs) | 46.4 ± 10.1 | 46.5 ± 9.8 | 46.3 ± 10.4 |
| Gender (Women/Men) | 79:18 | 42:6 | 37:12 |
| **Primary Outcomes** |  |  |  |
| BW (kg) | 103.1 ± 21.0 | 101.4 ± 21.6 | 104.7 ± 20.5 |
| FPG (mmol/L) | 6.2 ± 0.6 | 6.3 ± 0.7 | 6.2 ± 0.6 |
| **Secondary Outcomes** |  |  |  |
| <u>Anthropometry</u> |  |  |  |
| Height (m) | 1.67 ± 0.08 | 1.65 ± 0.08 | 1.68 ± 0.08 |
| BMI (kg/m<sup>2</sup>) | 37.1 ± 6.9 | 37.2 ± 7.4 | 37.0 ± 6.6 |
| <u>Blood pressure</u> |  |  |  |
| SBP (mmHg) | 133 ± 15 | 134 ± 17 | 132 ± 13 |
| DBP (mmHg) | 81 ± 8 | 80 ± 9 | 82 ± 8 |
| <u>Body composition</u> |  |  |  |
| TBFm (kg) | 47.2 ± 14.9 | 47.1 ± 15.0 | 47.3 ± 14.9 |
| TBFp (%) | 45.3 ± 7.1 | 45.9 ± 7.0 | 44.7 ± 7.3 |
| TBFFm (kg) | 55.3 ± 10.1 | 53.7 ± 9.9 | 57.0 ± 10.1 |
| AATm (kg) | 4.9 ± 1.9 | 4.8 ± 1.9 | 5.0 ± 1.9 |
| AATp (%) | 54.6 ± 8.0 | 54.4 ± 8.2 | 54.7 ± 7.9 |
| VATm (kg)<sup>#</sup> | 1.4 ± 0.7 | 1.3 ± 0.6 | 1.4 ± 0.8 |
| VATp (%)<sup>#</sup> | 30.0 ± 11.9 | 29.3 ± 11.5 | 30.6 ± 12.4 |
| SATm (kg)<sup>#</sup> | 3.3 ± 1.4 | 3.3 ± 1.5 | 3.4 ± 1.4 |
| SATp (%)<sup>#</sup> | 70.0 ± 11.9 | 70.7 ± 11.5 | 69.4 ± 12.4 |
| <u>Blood biochemistry</u> |  |  |  |
| HbA<sub>1c</sub> (mmol/mol) | 40.7 ± 4.9 | 41.2 ± 5.0 | 40.2 ± 4.7 |
| Insulin (μIU/mL) | 24.5 ± 14.9 | 23.7 ± 15.9 | 25.3 ± 13.9 |
| C-peptide (ng/mL) | 3.7 ± 1.2 | 3.7 ± 1.3 | 3.7 ± 1.1 |
| HOMA2-IR | 3.2 ± 1.6 | 3.0 ± 1.6 | 3.3 ± 1.6 |
| HOMA2-B | 132.9 ± 45.4 | 126.7 ± 43.3 | 138.9 ± 46.9 |
| HOMA2-S | 42.2 ± 26.6 | 43.7 ± 25.9 | 40.7 ± 27.4 |
| TC (mmol/L) | 5.1 ± 1.1 | 5.0 ± 1.1 | 5.1 ± 1.1 |
| TAG (mmol/L) | 1.5 ± 0.6 | 1.5 ± 0.7 | 1.4 ± 0.5 |
| HDL-C (mmol/L) | 1.2 ± 0.2 | 1.3 ± 0.3 | 1.2 ± 0.2 |
| LDL-C (mmol/L) | 3.2 ± 0.9 | 3.2 ± 0.9 | 3.3 ± 1.0 |
| ALT (IU/L) | 24.6 ± 15.6 | 26.0 ± 19.7 | 23.3 ± 10.1 |
| AST (IU/L) | 24.3 ± 10.8 | 25.0 ± 13.5 | 23.6 ± 7.4 |
| ALP (IU/L) | 80.2 ± 21.7 | 77.6 ± 19.8 | 82.7 ± 23.3 |
| GGT (IU/L) | 30.9 ± 21.4 | 27.5 ± 12.1 | 34.3 ± 27.4 |

Except gender, data are presented as mean ± SD. <sup>#</sup>n=92 participants at M0/CID1 (Fx: n=46, Placebo: n=46). ITT: intention-to-treat, M: month, CID: clinical investigation day, Fx: whole fruit feijoa powder, BW: body weight, FPG: fasting plasma glucose, BMI: body mass index, WC: waist circumference, SBP: systolic blood pressure, DBP: diastolic blood pressure, TBFm: total body fat mass, TBFp: total body fat percentage, TBFFm: total body fat-free mass, AATm: abdominal adipose tissue mass, AATp: abdominal adipose tissue percentage, VATm: visceral adipose tissue mass, VATp: visceral adipose tissue percentage, SATm: subcutaneous adipose tissue mass, SATp: subcutaneous adipose tissue percentage, HbA<sub>1c</sub>: haemoglobin A<sub>1c</sub>, HOMA2-IR: homeostatic model of insulin resistance 2, HOMA2-B: homeostatic model of β-cell function 2, HOMA2-S: homeostatic model of insulin sensitivity 2, TC: total cholesterol, TAG: triglycerides, HDL-C: high density lipoprotein cholesterol, LDL-C: low density lipoprotein cholesterol, ALT: alanine aminotransferase, AST: aspartate aminotransferase, ALP: alkaline phosphatase, GGT: gamma-glutamyl transferase.

**Figure 2:**
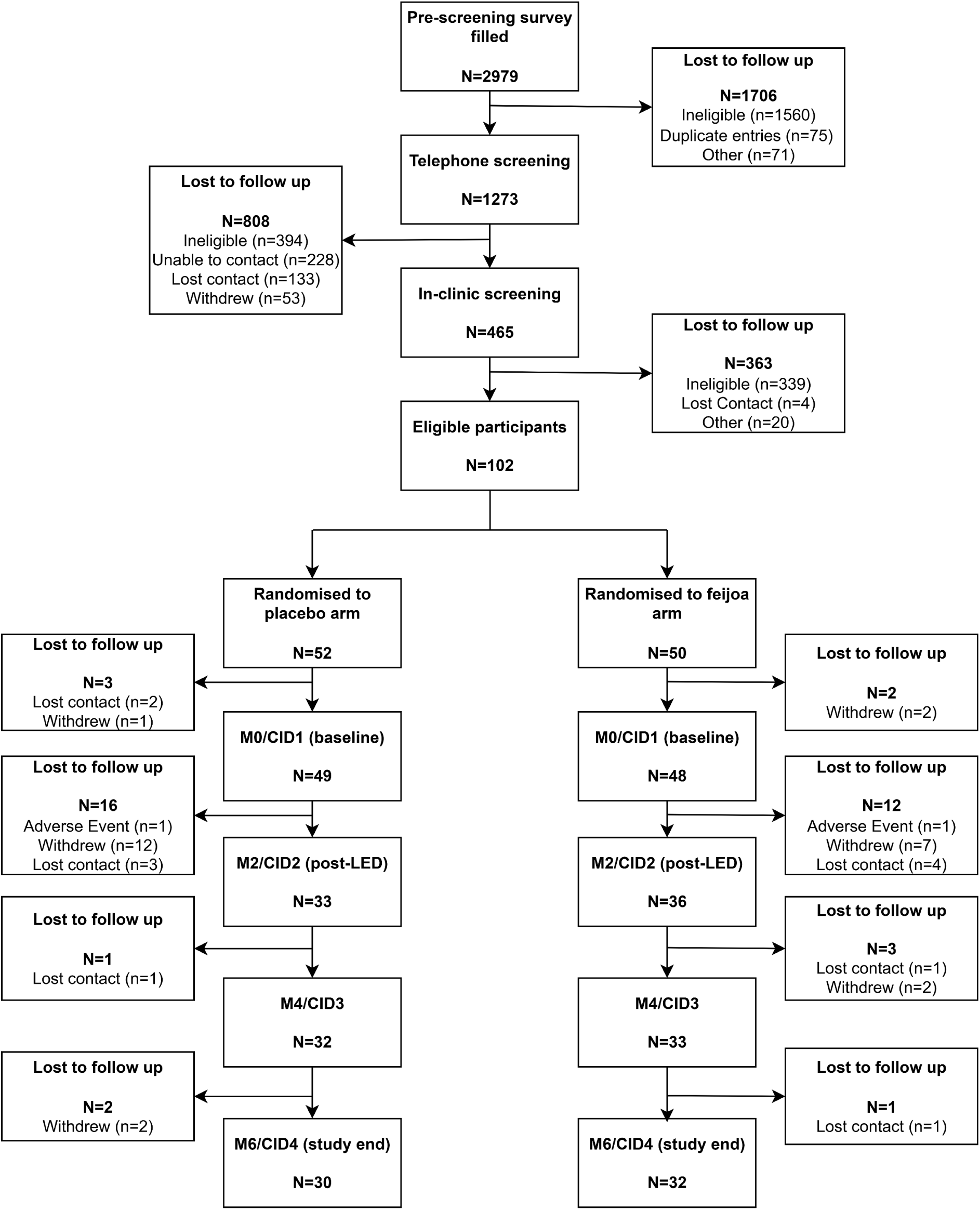
CONSORT flowchart describing progress of participants through the study. M: month, CID: clinical investigation day, LED: low energy diet, CONSORT: Consolidated Standards of Reporting Trials.

### Primary outcomes – BW, FPG (all and sub-cohort)

#### BW

BW was closely matched between treatment groups at baseline M0/CID1, with mean BMI above the cut point for obesity in both groups (Fx: 37.2 kg/m^2^; Placebo: 37.1 kg/m^2^). In the ITT cohort, no significant between-treatment difference in trajectory from baseline was observed (treatment×time_0-6m_: P=0.74). Fx treatment did not result in significantly greater decrease in BW vs. Placebo over 6 months. There was a significant change in BW over the study period, independent of treatment group (time_0-6m_: P<0.001), with a mean decrease of 7.2 kg during the 2 month LED phase in all participants (P<0.001). Whilst an increase was observed in all participants during the weight maintenance phase (ΔBW_2-6m_: 2.1 kg; P<0.001), BW remained lower than baseline at study end (ΔBW_0-6m_: -5.1 kg; P<0.001). BMI followed the same trajectory from baseline and between treatment groups.

#### FPG, all, n=97

In the full ITT cohort (n=97), there was a significant difference in trajectory from baseline between the two treatment groups (treatment×time_0-6m_: P=0.05). However, Fx was not significantly lower than Placebo at any timepoint. In addition, and importantly, sensitivity analysis showed that removal of the single extreme FPG outlier from the Placebo group at the end of intervention timepoint (M6/CID4, FPG: 9.6 mmol/L) resulted in loss of significance (Figure 3A, treatment×time_0-6m_: P=0.09). There was a significant change in FPG between baseline and M6/CID4, independent of treatment group (time_0-6m_: P<0.001). In line with BW, *post hoc* analysis revealed that FPG in all participants decreased during the LED weight loss phase (ΔFPG_0-2m_: -0.5 mmol/L; P<0.001). Despite an increase during the weight maintenance phase (ΔFPG_2-6m_: 0.3 mmol/L; P<0.001), FPG in all participants remained significantly lower than baseline at all timepoints (P<0.001, all).

**Figure 3:**
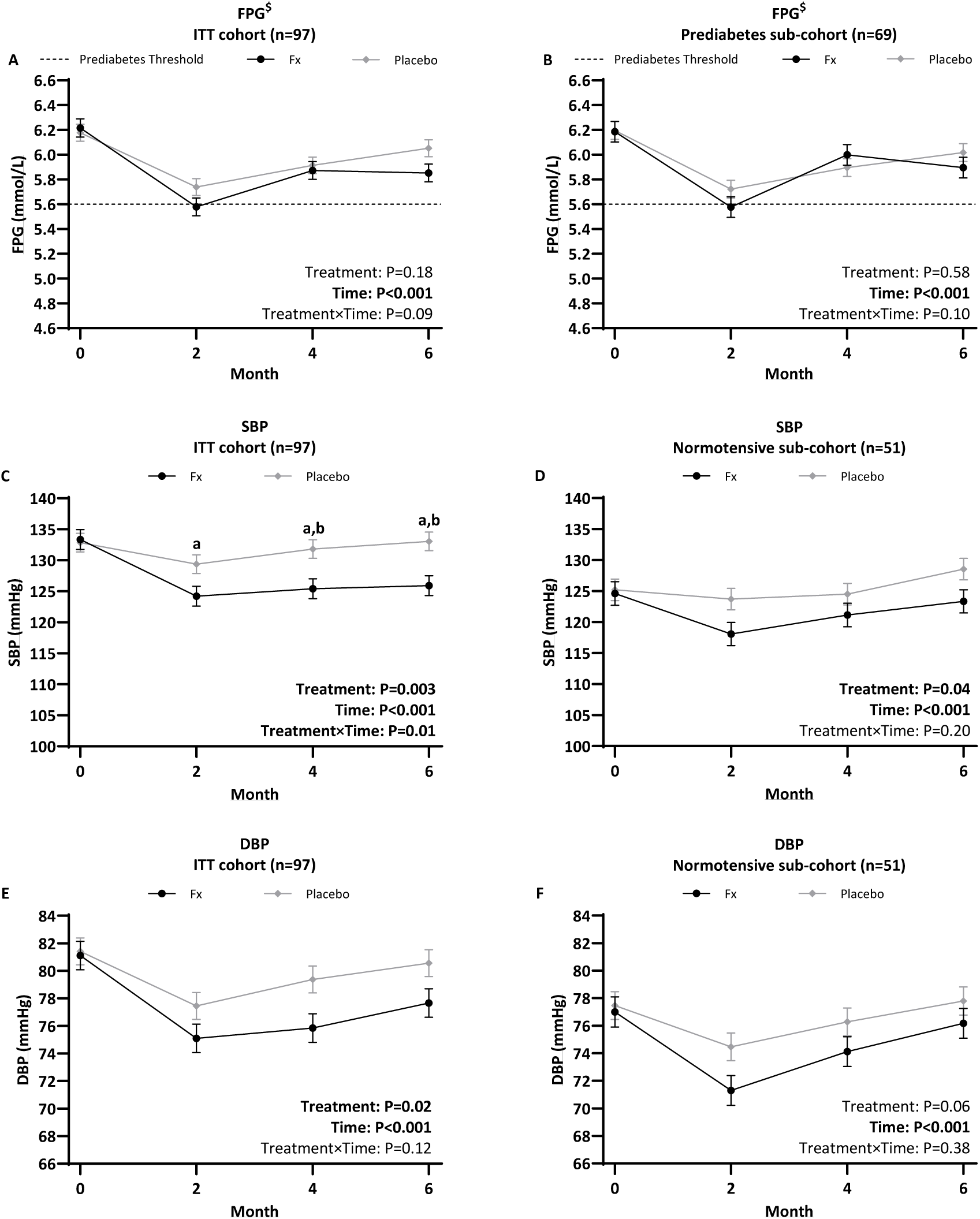
Linear mixed-effects model for FPG, SBP, and DBP in the ITT cohort (A, C & E), and sub-cohorts with prediabetes (B) and normotension (D & F) confirmed at CID1/baseline. Data is presented as estimated marginal mean ± SEM. ^$^Extreme outlier removed prior to analysis (n=1, Placebo, M6/CID4, FPG: 9.6 mmol/L). Post hoc differences shown as following superscripts; ^a^P≤0.05 compared to M0/CID1 in Fx group, ^b^P≤0.05 between treatments at same timepoint. ITT: intention-to-treat, M: month, CID: clinical investigation day, SEM: standard error of the mean, BW: body weight, FPG: fasting plasma glucose, Fx: whole fruit feijoa powder, SBP: systolic blood pressure, DBP: diastolic blood pressure.

#### FPG, sub-cohort (prediabetes only), n=69

The mean ± SD delay between in-clinic screening/enrolment and baseline assessment was 33 ± 8 days, during which FPG in 16 participants lowered to normoglycaemia (FPG ≤5.5 mmol/L, Fx: n=8, Placebo: n=8) and 12 participants worsened to T2D (FPG ≥7.0 mmol/L, Fx: n=9, Placebo: n=3). Analyses of the sub-cohort with confirmed prediabetes at baseline (n=69, extreme outlier removed) showed that no significant difference in change in FPG over time was apparent between the treatment groups (treatment×time_0-6m_: P=0.10).

Sensitivity analysis showed that inclusion of the single extreme outlier datapoint at the M6/CID4 timepoint in this sub-cohort (n=69) did not change this outcome. The significant main effect of time on FPG independent of treatment group persisted (Figure 3B, time_0-6m_: P<0.001). *Post hoc* analyses showed a significant decrease in FPG during the LED weight loss phase in all participants, with FPG remaining lower than baseline at all timepoints (P<0.001, all).

### Secondary outcomes

There was a significant change between baseline and M6/CID4, independent of treatment group, in all other anthropometry, body composition, and blood biochemistry outcomes (Table 2, time_0-6m_: P<0.01, all) except SATp and the liver enzyme ALP, with consistent improvements during the LED weight loss phase. There were no significant effects of Fx vs. Placebo treatment over the 6 month intervention for any of these secondary variables (Table 2, treatment×time_0-6m_: P>0.05, all).

**Table 2:** Primary and secondary outcomes in the ITT cohort (n=97).

|  | Fx (n=48) |  |  |  | Placebo (n=49) |  |  |  | P value |  |  |
| --- | --- | --- | --- | --- | --- | --- | --- | --- | --- | --- | --- |
|  | M0/CID1 | M2/CID2 | M4/CID3 | M6/CID4 | M0/CID1 | M2/CID2 | M4/CID3 | M6/CID4 | Treat | Time | Treat×Time |
| **Primary Outcomes** |  |  |  |  |  |  |  |  |  |  |  |
| BW (kg) | 103.1 (0.9) | 96.0 (0.9) | 97.2 (0.9) | 98.5 (0.9) | 103.2 (0.8) | 95.9 (0.8) | 96.7 (0.8) | 97.6 (0.8) | 0.68 | <0.001 | 0.74 |
| FPG (mmol/L)<sup>§</sup> | 6.2 (0.1) | 5.6 (0.1) | 5.9 (0.1) | 5.9 (0.1) | 6.2 (0.1) | 5.7 (0.1) | 5.9 (0.1) | 6.1 (0.1) | 0.18 | <0.001 | 0.09 |
| **Secondary Outcomes** |  |  |  |  |  |  |  |  |  |  |  |
| <u>**Anthropometry**</u> |  |  |  |  |  |  |  |  |  |  |  |
| BMI (kg/m<sup>2</sup>) | 37.2 (0.3) | 34.5 (0.3) | 34.9 (0.3) | 35.4 (0.3) | 37.1 (0.3) | 34.5 (0.3) | 34.8 (0.3) | 35.1 (0.3) | 0.72 | <0.001 | 0.84 |
| WC (cm) | 114.1 (0.9) | 107.9 (0.9) | 108.5 (0.9) | 110.4 (0.9) | 114.0 (0.8) | 106.9 (0.8) | 107.9 (0.8) | 109.3 (0.8) | 0.42 | <0.001 | 0.74 |
| <u>**Blood pressure**</u> |  |  |  |  |  |  |  |  |  |  |  |
| SBP (mmHg) | 133 (2) | 124 (2)<sup>a</sup> | 125 (2)<sup>a,b</sup> | 126 (2)<sup>a,b</sup> | 133 (2) | 129 (2) | 132 (2)<sup>b</sup> | 133 (2)<sup>b</sup> | **0.003** | <0.001 | **0.01** |
| DBP (mmHg) | 81 (1) | 75 (1) | 76 (1) | 78 (1) | 81 (1) | 77 (1) | 79 (1) | 81 (1) | **0.02** | <0.001 | 0.12 |
| <u>**Body composition**</u> |  |  |  |  |  |  |  |  |  |  |  |
| TBFm (kg) | 47.0 (0.7) | 42.0 (0.7) | 42.3 (0.7) | 43.3 (0.7) | 47.1 (0.6) | 41.9 (0.6) | 41.7 (0.6) | 42.4 (0.6) | 0.60 | <0.001 | 0.54 |
| TBFp (%) | 45.1 (0.4) | 43.0 (0.4) | 42.9 (0.4) | 43.5 (0.4) | 45.2 (0.3) | 43.2 (0.3) | 42.6 (0.3) | 42.9 (0.3) | 0.67 | <0.001 | 0.35 |
| TBFFm (kg) | 55.5 (0.3) | 53.6 (0.3) | 54.3 (0.3) | 54.6 (0.3) | 55.5 (0.3) | 53.3 (0.3) | 54.2 (0.3) | 54.6 (0.3) | 0.69 | <0.001 | 0.82 |
| AATm (kg) | 4.9 (0.1) | 4.2 (0.1) | 4.2 (0.1) | 4.4 (0.1) | 4.9 (0.1) | 4.1 (0.1) | 4.1 (0.1) | 4.2 (0.1) | 0.48 | <0.001 | 0.55 |
| AATp (%) | 54.2 (0.5) | 51.4 (0.5) | 51.6 (0.5) | 51.9 (0.5) | 54.3 (0.5) | 51.2 (0.5) | 50.7 (0.5) | 51.0 (0.5) | 0.40 | <0.001 | 0.33 |
| VATm (kg)<sup>#</sup> | 1.9 (0.1) | 1.6 (0.1) | 1.6 (0.1) | 1.6 (0.1) | 1.9 (0.0) | 1.5 (0.0) | 1.5 (0.0) | 1.6 (0.0) | 0.45 | <0.001 | 0.35 |
| VATp (%)<sup>#</sup> | 40.4 (0.7) | 39.5 (0.7) | 39.3 (0.7) | 39.0 (0.7) | 40.6 (0.7) | 38.2 (0.7) | 38.8 (0.7) | 38.6 (0.7) | 0.43 | **0.003** | 0.64 |
| SATm (kg)<sup>#</sup> | 3.3 (0.1) | 2.9 (0.1) | 2.9 (0.1) | 3.0 (0.1) | 3.3 (0.1) | 2.8 (0.1) | 2.8 (0.1) | 2.9 (0.1) | 0.53 | <0.001 | 0.82 |
| SATp (%)<sup>#</sup> | 70.2 (0.7) | 69.3 (0.7) | 69.9 (0.7) | 70.7 (0.7) | 70.1 (0.7) | 70.5 (0.7) | 69.9 (0.7) | 70.4 (0.7) | 0.70 | 0.73 | 0.61 |
| <u>**Blood biochemistry**</u> |  |  |  |  |  |  |  |  |  |  |  |
| HbA<sub>1c</sub> (mmol/mol) | 40.8 (0.5) | 38.2 (0.5) | 39.1 (0.5) | 39.6 (0.5) | 40.5 (0.4) | 38.2 (0.4) | 39.0 (0.4) | 39.5 (0.4) | 0.79 | <0.001 | 0.95 |
| Insulin (μIU/mL) | 20.3 (1.1) | 12.6 (1.1) | 15.6 (1.1) | 16.3 (1.1) | 20.7 (1.1) | 13.8 (1.1) | 15.9 (1.1) | 17.4 (1.1) | 0.39 | <0.001 | 0.79 |
| C-peptide (ng/mL) | 3.6 (0.1) | 2.9 (0.1) | 3.0 (0.1) | 3.1 (0.1) | 3.7 (0.1) | 3.1 (0.1) | 3.1 (0.1) | 3.3 (0.1) | 0.20 | <0.001 | 0.57 |
| HOMA2-IR | 3.1 (0.1) | 2.1 (0.1) | 2.4 (0.1) | 2.5 (0.1) | 3.1 (0.1) | 2.2 (0.1) | 2.4 (0.1) | 2.7 (0.1) | 0.42 | <0.001 | 0.85 |
| HOMA2-B | 130.6 (3.7) | 119.2 (3.7) | 123.4 (3.7) | 128.2 (3.7) | 132.3 (3.5) | 116.3 (3.5) | 119.3 (3.5) | 121.9 (3.5) | 0.42 | <0.001 | 0.45 |
| HOMA2-S | 37.6 (1.1) | 59.7 (1.1) | 48.5 (1.1) | 46.5 (1.1) | 36.8 (1.1) | 55.4 (1.1) | 47.8 (1.1) | 43.4 (1.1) | 0.42 | <0.001 | 0.81 |
| TC (mmol/L) | 5.1 (0.1) | 4.5 (0.1) | 5.2 (0.1) | 5.1 (0.1) | 5.1 (0.1) | 4.4 (0.1) | 5.1 (0.1) | 5.2 (0.1) | 0.63 | <0.001 | 0.43 |
| TAG (mmol/L) | 1.3 (1.0) | 1.2 (1.0) | 1.3 (1.0) | 1.3 (1.0) | 1.4 (1.0) | 1.2 (1.0) | 1.3 (1.0) | 1.4 (1.0) | 0.75 | <0.001 | 0.34 |
| HDL-C (mmol/L) | 1.3 (0.0) | 1.2 (0.0) | 1.3 (0.0) | 1.4 (0.0) | 1.3 (0.0) | 1.2 (0.0) | 1.3 (0.0) | 1.3 (0.0) | 0.63 | <0.001 | 0.33 |
| LDL-C (mmol/L) | 3.2 (0.1) | 2.7 (0.1) | 3.3 (0.1) | 3.2 (0.1) | 3.2 (0.1) | 2.6 (0.1) | 3.2 (0.1) | 3.2 (0.1) | 0.70 | <0.001 | 0.22 |
| ALT (IU/L) | 21.9 (1.1) | 23.9 (1.1) | 18.5 (1.1) | 17.8 (1.1) | 21.8 (1.0) | 23.8 (1.0) | 18.7 (1.0) | 18.5 (1.0) | 0.83 | <0.001 | 0.93 |
| AST (IU/L) | 23.1 (1.0) | 24.1 (1.0) | 20.2 (1.0) | 20.7 (1.0) | 23.0 (1.0) | 25.9 (1.0) | 21.4 (1.0) | 21.9 (1.0) | 0.16 | <0.001 | 0.52 |
| ALP (IU/L) | 80.5 (1.4) | 78.8 (1.4) | 80.5 (1.4) | 78.6 (1.4) | 80.7 (1.3) | 77.4 (1.3) | 79.6 (1.3) | 79.9 (1.3) | 0.90 | 0.07 | 0.53 |
| GGT (IU/L) | 26.5 (1.0) | 22.8 (1.0) | 23.8 (1.0) | 23.5 (1.0) | 26.7 (1.0) | 22.6 (1.0) | 23.7 (1.0) | 23.4 (1.0) | 0.95 | <0.001 | 0.99 |

#### SBP, DBP

Notably, however, there was a significant treatment×time interaction for SBP (Figure 3C, treatment×time_0-6m_: P=0.01), reflecting the difference in trajectory from baseline of SBP between treatment groups over 6 months*. Post hoc* analyses showed SBP to be significantly lower in Fx treatment compared to Placebo at M4/CID3 and M6/CID4 (P≤0.05, both). Fx treatment resulted in significantly greater improvement in SBP vs. Placebo over 6 months. There was no significant interaction for DBP over the 6 month intervention (Figure 3E, treatment×time_0-6m_: P=0.12).

#### BP, sub-cohort (normotension only)

47% of participants (Fx, n=24; Placebo, n=22) had hypertension at baseline. In the sub-cohort of 51 normotensive participants, there was no significant interaction and hence no significant difference in the change in trajectory of SBP or DBP between treatment groups over 6 months (treatment×time_0-6m_, SBP: P=0.20, DBP: P=0.38). Contrary to the ITT cohort, Fx did not result in significantly greater decrease in SBP over 6 months In this normotensive sub-cohort. A significant change in SBP and DBP over 6 months, independent of treatment group, persisted (Figure 3D & 3F, time_0-6m_: P<0.001, both).

### Raw data (no imputation)

Analyses from all observed data (i.e. without imputation) were consistent with ITT analyses across all primary and secondary outcomes (results not shown).

## Discussion

The FERDINAND RCT investigated the effects of Fx supplementation on BW and T2D biomarkers in a multi-ethnic cohort of adults living with overweight and obesity, during and following 8 weeks of LED-induced weight loss. Participants were enrolled into the intervention with overweight and dysglycaemia, and with a desire to achieve and maintain significant weight loss of ∼8-10 %. The study comprised an acute 2-month phase of LED meal replacement-driven weight loss, followed by a 4-month phase of healthy eating lifestyle advice aiming to maintain the weight lost. Compliance to the meal replacements and intervention powder demonstrated the successful completion of the treatments over the study period.

Regarding the co-primary outcomes, when analysed independent of treatment group, BW changed significantly over the 6 months as expected, with a post-LED decrease over 2 months followed by rebound during the weight maintenance phase. BW was not significantly different between the treatments throughout the study, with no evidence of greater weight loss in the Fx treatment group. The hypothesised enhancement of Fx on weight loss was grounded on proposed anti-obesity mechanisms attributed to polyphenols. Experimental evidence suggests that polyphenols may inhibit adipocyte differentiation (64), increase fatty acid oxidation (65), and modulate gut microbiota composition in ways that favour improved metabolic regulation (66,67). However, translation of these mechanistic insights into clinically meaningful weight loss in humans has been inconsistent. A recent meta-analysis of 40 RCTs administering polyphenol supplementation (31-3000 mg/day for 1-12 months) reported BW loss of 0.36 kg (95 % CI: -0.68 to -0.05; P=0.02) (68). Although statistically significant, the magnitude of weight loss due to supplementation alone is unlikely to be clinically meaningful, particularly when contrasted with the ≥5 % BW loss commonly used to define clinical relevance (69). Furthermore, the substantial energy deficit imposed during the LED phase of FERDINAND likely exerted a dominant physiological effect on BW. It is therefore plausible that any incremental influence of polyphenols on adiposity was comparatively small and effectively masked by the magnitude of LED-induced weight loss. This interpretation is consistent with previous trials that have combined polyphenol supplementation with an LED, which similarly reported no additional effects on BW or body composition (70–73).

FPG followed the weight loss trajectory and decreased and then rebounded post-LED in both treatment groups. Whilst borderline significant improvement in FPG was observed in the Fx vs. Placebo group, this effect was lost when sensitivity analysis was undertaken. This highlights that the effect of Fx on FPG was statistically weak and influenced by a single outlier point in the Placebo group at the end of study assessment. Furthermore, these observations included 28 participants in whom FPG either reverted to normoglycaemia or progressed to T2D between the screening visit and start of the intervention, i.e. were no longer prediabetic. Removal of these participants from the analysis eliminated the borderline difference between the two treatments, irrespective of the single extreme outlier, but notably also decreased power to detect any change as statistically significant.

Nevertheless, sustained lowering of FPG in the Fx group is plausible when considering the bioactive profile of the whole fruit feijoa powder. Approximately 60 % of the powder’s known polyphenolic content consisted of catechins (Anagenix Ltd, personal communication, March 19 2026). A recent meta-analysis by Wan and colleagues, encompassing 28 RCTs administering catechin supplementation (63-1344 mg/day for 1-12 months), reported a mean FPG lowering of 0.10 mmol/L (95 % CI: -0.18 to - 0.03; P<0.001) (74). Although FPG did not differ significantly between treatment arms at study end in FERDINAND (-0.13 mmol/L, 95 % CI: -0.49 to 0.23), the model-estimated differences were numerically comparable to the pooled effect size reported in the meta-analysis (74). At first glance, this comparison appears incongruent given the markedly lower known catechin dose delivered in FERDINAND (0.60 mg/day), which was over 100-fold lower than the lowest dose included in Wan and colleagues’ meta-analysis (63 mg/day). In addition, FERDINAND incorporated a structured LED regimen, whereas only two trials within Wan and colleagues’ meta-analysis included an LED component (70,72). It could be argued that relative to the studies included in Wan and colleagues’ meta-analysis, the FPG findings in FERDINAND were primarily driven by the structured LED rather than by polyphenol exposure. However, comparison with the two trials within the meta-analysis that also incorporated LEDs does not fully support this assertion. Despite administering catechin doses that were orders of magnitude higher than those used in FERDINAND (1125 mg/day and 300 mg/day), the magnitude of FPG change reported in these trials was comparable to that observed in FERDINAND. Furthermore, the whole fruit feijoa powder contained a matrix of bioactive compounds, including procyanidins and ellagic acid, which may exert additive or synergistic metabolic effects. *In vitro* and animal studies suggest that combinations of polyphenols can produce synergistic improvements in inflammation (75) and oxidative stress (76,77), mechanisms closely linked to glycaemic regulation.

Additionally, the potential role of ABA in the sustained FPG lowering in the Fx group must also be considered. The 1150 mg of whole fruit feijoa powder used in the FERDINAND study provided 54 μg/day of ABA (Anagenix Ltd, personal communication, March 19 2026). RCTs investigating daily ABA supplementation have utilised a dose of 55 μg/day of ABA for 75 days in healthy adults (n=10) (46), and alternatively, 2 g/day of dwarf peach powder, known to contain ABA, over a 3-month period in individuals with prediabetes (n=65) (78). While the ABA dosage was not explicitly reported in the latter trial (78), its use of a fruit-derived ABA source in a population with prediabetes makes it one of the few studies to examine such an intervention in this at-risk group, and thus a particularly relevant comparator to FERDINAND. Despite these differences, both studies reported improvements in fasting glycaemia by the end of the supplementation period (46,78).

Amongst the secondary outcomes, only SBP in the Fx group significantly improved compared to Placebo over the 6 month intervention. Since there was no BP exclusion criteria for the trial, a large number of participants with overweight and associated biomarkers of poor metabolic health were enrolled with previously undiagnosed hypertension at baseline. When these participants were removed from the data analyses, the significant effect of Fx vs. Placebo over 6 months was lost. Whilst this may be in part due to the decrease in sample size, it is possible that the observed differences in SBP between the treatments may be driven by the sub-cohort of participants with uncontrolled and/or controlled hypertension.

From a mechanistic perspective, flavonoids such as catechins, the most abundant polyphenolic class in whole fruit feijoa powder, have established vasoprotective properties. Flavonoids have been shown to enhance endothelial nitric oxide bioavailability, inhibit angiotensin-converting enzyme activity, and attenuate oxidative stress (79). Consistent with these mechanisms, a recent meta-analysis of 31 RCTs implementing catechin supplementation (20-1344 mg/day for 2-24 weeks) reported a mean SBP lowering of 1.56 mmHg (95 % CI: -2.75 to -0.37; P<0.001) (74). Notably, the model-estimated mean SBP lowering observed in FERDINAND (-5.93 mmHg, 95 % CI: -15.42 to 3.56) was numerically greater than that reported in 28 of the 31 trials included in this meta-analysis (74), despite the substantially lower catechin dose administered. Furthermore, when compared to the only trial within the meta-analysis that incorporated an LED (70), the magnitude of SBP lowering in FERDINAND was comparable. One potential explanation for these findings is the combined influence of LED-induced metabolic improvements and the polyphenolic profile of whole fruit feijoa. Experimental evidence suggests that polyphenolic compounds may exert synergistic vascular effects. For example, Zhang and colleagues demonstrated that combined curcumin and luteolin exposure produced greater inhibition of TNF-α-induced vascular inflammation than either compound alone in both human and murine models (80). It is therefore plausible that the polyphenolic matrix of whole fruit feijoa contributed to vascular benefits beyond what might be expected from catechin content alone.

As noted previously, the intervention conducted by Taghavi and colleagues in Iran remains the only prior clinical trial to investigate whole fruit feijoa supplementation in humans (58). Whilst the FERDINAND study is the first to investigate supplementation with a whole fruit feijoa powder for T2D prevention, it was not without some limitations. Owing to a number of challenges in recruiting participants into this study during the COVID pandemic, underpinned by many individuals with prediabetes being unaware of their adverse glycaemic status (81), and in the absence of validated predictive tools to pinpoint individuals with prediabetes amongst many with overweight yet normoglycaemia (82), the sample size was below the *a priori* target. Hence the study may not have detected small yet clinically important differences in FPG between treatments. Additionally, a sub-cohort of participants reverted to normoglycaemia or progressed to T2D between enrolment and baseline assessment, with the average time between these visits being 33 days. Hence the cohort with confirmed prediabetes at baseline was smaller than planned. Regarding attrition, a 20 % dropout rate is typically expected in LED and VLED interventions (15). In contrast, the dropout rate in FERDINAND was 36 %, of which 4 out of 5 of these participants withdrew during the 2 month LED. It must be considered that FERDINAND implemented a strict total meal replacement strategy, including restrictions to non-caloric beverage consumption, which may have contributed to the higher than expected dropout rate. Nevertheless, results that were generated from observed data (i.e. no imputation) were consistent with ITT analyses, further supporting the use of LVCF for imputing missing data.

In conclusion, despite no significant effect on BW or FPG beyond that achieved through a commercial LED alone, the FERDINAND study provides evidence to support Fx supplementation during weight loss for improvement in SBP in adults with overweight plus current/recent history of prediabetes. Further studies in larger cohorts are warranted to fully evaluate the contribution that whole fruit feijoa powder may make to improved blood pressure.

## Data Availability

The data produced in the present study may be available upon reasonable request to the corresponding author. However, individual participant data will not be made available. This is in accordance with Southern Health and Disabilities Ethics Committee (New Zealand) application that all data generated will only be used for this study.

## Declarations

### Ethics statement

Ethics approval was received from the New Zealand Health and Disability Ethics Committee (2022 EXP 12032) and prospectively registered with the Australian New Zealand Clinical Trials Registry (ACTRN: 12622000210774). Study participants were provided with all study details, explained to them in writing and in person before they provided written informed consent.

### Data availability statement

The datasets generated from this study will be restricted in line with ethical approvals. Requests to access the datasets should be directed to Jennifer Miles-Chan. Individual participant data will not be made available. This is in accordance with National Health and Disability Ethics Committees application that all data generated will only be used for the purpose of this study.

### Author contributions

IM: Investigation, Project administration, Statistical analysis, Writing – original draft. IS-B: Conceptualisation, Methodology, Supervision, Statistical analysis, Writing – review & editing. JL: Investigation, Writing – review & editing. LP: Conceptualisation, Methodology, Supervision, Writing – review & editing. RM: Conceptualisation, Supervision, Writing – review & editing. SP: Conceptualisation, Funding acquisition, Methodology, Supervision, Writing – review & editing. JM-C: Conceptualisation, Funding acquisition, Methodology, Supervision, Writing – review & editing.

## Acknowledgements

We thank Dr William Zhu for collection and processing of blood samples; Dr Shakeela Jayasinghe for conducting and maintaining confidentiality of intervention blinding; Registered Dietitians Dr Amy Liu and Shivaanee Ram for reviewing the diet design and administering group dietary consultation meetings with participants; Christine Keven at the Analytical and Molecular Research Platform, Liggins Institute, for blood biochemical measurements; Associate Professor Beatrix Jones and Jessica McLay at the Department of Statistics, University of Auckland, for their statistical advice. We also appreciate the participants who took part in this study.

